# Multimodal Electronic Health Record Foundation Models with Electrocardiogram for Cardiovascular Disease Prediction

**DOI:** 10.1101/2025.11.10.25339886

**Authors:** Junmo Kim, Young-Kwan Kim, Kwangsoo Kim

## Abstract

Electronic health record (EHR) foundation models (FMs) have improved clinical task performance by learning comprehensive clinical context from sequential medical records called patient trajectories. However, existing multimodal approaches generate representations separately for each modality and integrate them only at the final prediction stage, without capturing temporal relationships across data types. Here, we propose TRIM, text-based representations for integrated multimodal trajectories, which converts unstructured data into text-based representations via their diagnostic reports and integrates them directly into patient trajectories. This enables EHR FMs to simultaneously analyze structured and unstructured longitudinal data while preserving temporal context. To develop and evaluate TRIM, we trained EHR FMs with electrocardiogram data to predict cardiovascular diseases (CVDs). TRIM-integrated EHR FMs improved overall CVD prediction performance. Model interpretation revealed that models prioritized ECG diagnostic information, focusing on clinically established risk factors. Survival analysis validated model decisions, with TRIM integration consistently increasing hazard ratios. TRIM provides a generalizable framework for integrating multimodal medical data into EHR FMs.

## Introduction

With the availability of large structured electronic health record (EHR) datasets^1,2^, EHR foundation models (FMs) have emerged to enhance the performance of diverse clinical tasks^3–6^. EHR FMs are trained on patient trajectories, which are patient-level sequential medical codes extracted from EHR databases. As each patient trajectory corresponds to a natural language sentence, EHR FMs have followed the paradigms of natural language processing. By utilizing entire patient medical histories rather than specific curated variables, EHR FMs learn comprehensive medical contexts to fulfill given clinical tasks, such as predicting patient outcomes^3,7^, simulating patient status^6^, and synthesizing EHR data^8^.

Despite current advances in EHR FMs, only a few studies have utilized multiple data modalities with EHR FMs. Mataraso et al. proposed Clinical and Omics Multimodal analysis Enhanced with Transfer learning (COMET)^9^. COMET complemented the data scarcity of omics data by integrating EHR FM representations that contain information from many more patients. Soenksen et al. proposed the Holistic Artificial Intelligence in Medicine (HAIM) framework to leverage multimodal inputs^10^. HAIM concatenated multimodal representations generated from respective pretrained models to construct final integrated inputs for clinical tasks. While many other studies have utilized structured EHR data with unstructured data, EHR FMs have been rarely used. Although COMET and HAIM achieved improved performance on diverse clinical tasks, they generated representations from EHR and non-EHR data separately and integrated them at the final stage. This approach does not capture the temporal relationships between medical codes and other modalities throughout the patient timeline.

In this study, we propose text-based representations for integrated multimodal trajectories (TRIM), to integrate unstructured data into patient trajectories for EHR FMs. The motivation behind TRIM is that most unstructured medical data, such as electrocardiogram (ECG), chest X-ray (CXR), and computed tomography (CT), are accompanied by textual reports written by machines or clinicians. Inspired by MedRep^11^, which provides a representation for each medical concept (code) using free-text descriptions, we integrated text-based representations of unstructured data into original patient trajectories by generating MedRep-style representations from the textual reports of the unstructured data. In this way, EHR FMs can simultaneously analyze the comprehensive context of structured and unstructured longitudinal data while considering time intervals between records.

To develop and evaluate TRIM, we trained EHR FMs with ECG data to predict the first occurrence of cardiovascular diseases (CVDs), including heart failure (HF), ischemic heart disease (IHD), coronary artery disease (CAD), and stroke, to contribute to the prevention and early-management of CVDs. Outcomes that can be directly diagnosed from ECGs (e.g., arrhythmia and atrial fibrillation) were excluded to effectively evaluate the impact of integrated unstructured data, thereby preventing the models from excessively relying on raw ECG signals. We used EHR and ECG data from one publicly available dataset, MIMIC-IV^12^, and one private dataset from Seoul National University Hospital, a tertiary hospital in the Republic of Korea. The results demonstrate that leveraging TRIM-style ECG records enhances CVD prediction performance and that EHR FMs focus more on integrated ECG data rather than original EHR records when making decisions.

## Results

In this study, we proposed a multimodal EHR FM framework to predict four CVDs (HF, IHD, CAD, and stroke) within one year after ECG measurement through a multilabel classification approach (Fig. 1a). We defined the prediction timepoint (index date) as the last ECG measurement before the first CVD occurrence for the CVD group, with a one-month washout period to exclude imminent events. For the non-CVD group, the prediction timepoint (index date) was the last ECG measurement, and patients were included only if they had at least one visit record more than one year later (follow-up period). TRIM representations were generated by LLM-enriched ECG diagnosis descriptions and the text-based representation generator of MedRep (Figs. 1b-1c). In the CVD prediction process (Fig. 1d), each ECG record was converted into a TRIM representation and integrated into the patient trajectory alongside EHR records. An EHR FM generated “trajectory representations” from these integrated trajectories, while an ECG encoder generated “ECG representations” from raw ECG signals of the last ECG measurement. These trajectory and ECG representations were processed through separate linear layers for independent and combined modalities, then concatenated and passed through a final prediction head.

**Fig 1.**
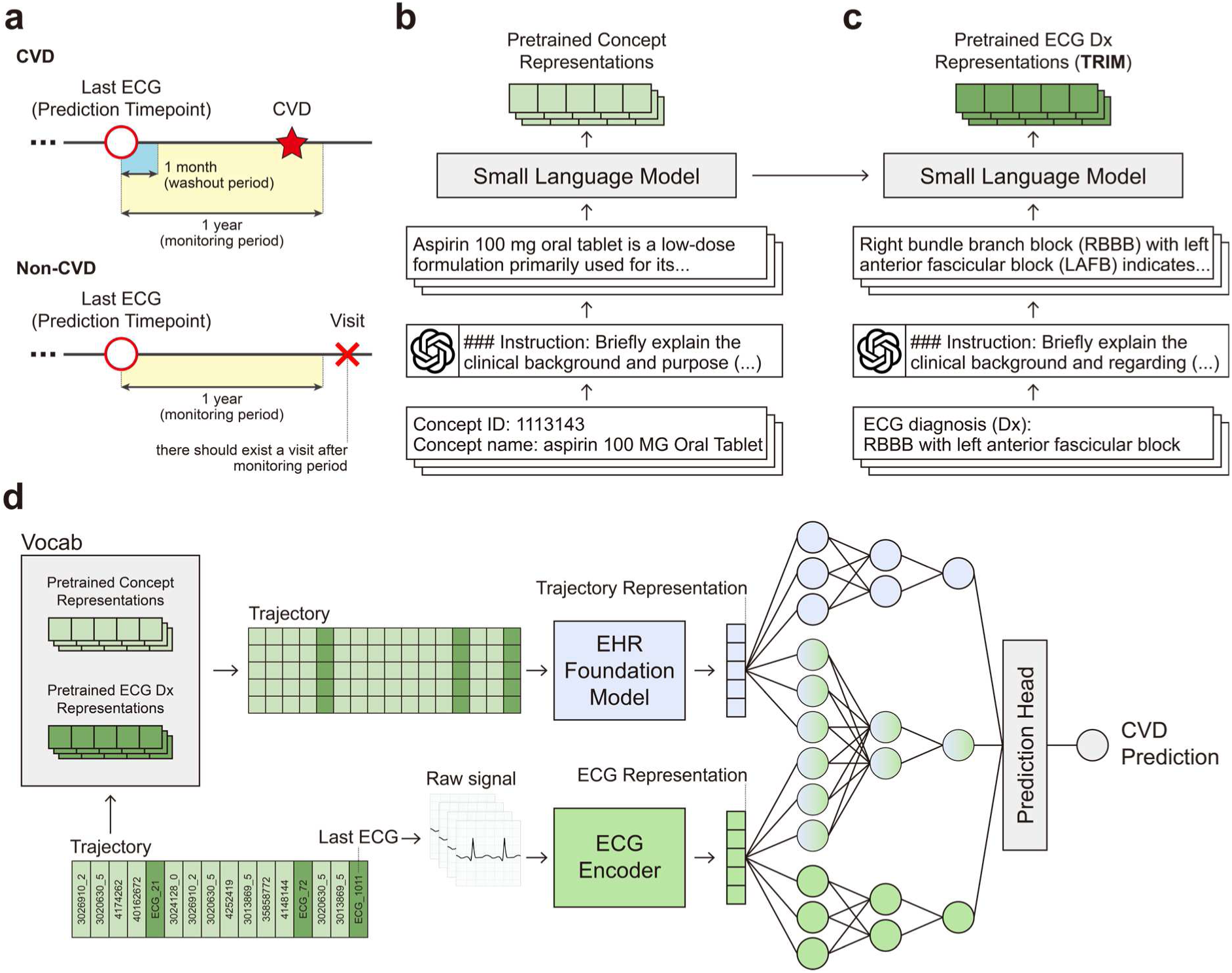
Implementation process for CVD prediction using multimodal EHR FMs with TRIM. **a,** Prediction timepoint definition for CVD and non-CVD groups. **b**, Text-based representation generation process of medical concepts in EHR. Each phrase-level concept name is enriched with LLM-generated descriptions. **c**, Text-based representation generation process of phrase-level ECG diagnoses (TRIM). TRIM representations were generated by LLM-enriched ECG diagnosis descriptions and a small language model trained on medical concept descriptions. **d**, CVD prediction process with representation-based EHR FMs, TRIM, and raw ECG signals. Representations generated by EHR FM and ECG encoder are integrated to predict CVDs.

### Baseline characteristics

For both MIMIC-IV and SNUH datasets, we used 85% of data for pretraining and 15% for finetuning (Table 1). Of the 257,992 patients in MIMIC-IV and 39,128 patients in SNUH, we allocated 219,293 and 33,258 patients to the pretraining dataset, and 38,699 and 5,870 patients to the finetuning dataset, respectively. Age, sex, and outcome distributions were significantly different between datasets (all p<0.0001). HF (7.37% vs.4.16%) and CAD (10.68% vs. 6.21%) incidence was higher in MIMIC-IV, while IHD (1.55% vs. 4.52%) and stroke (2.06% vs. 3.43%) was higher in SNUH. Although MIMIC-IV contained more patients overall, SNUH showed higher patient data density, with more visits, EHR records, and ECG records per patient, as well as more unique ECG diagnoses.

**Table 1.**
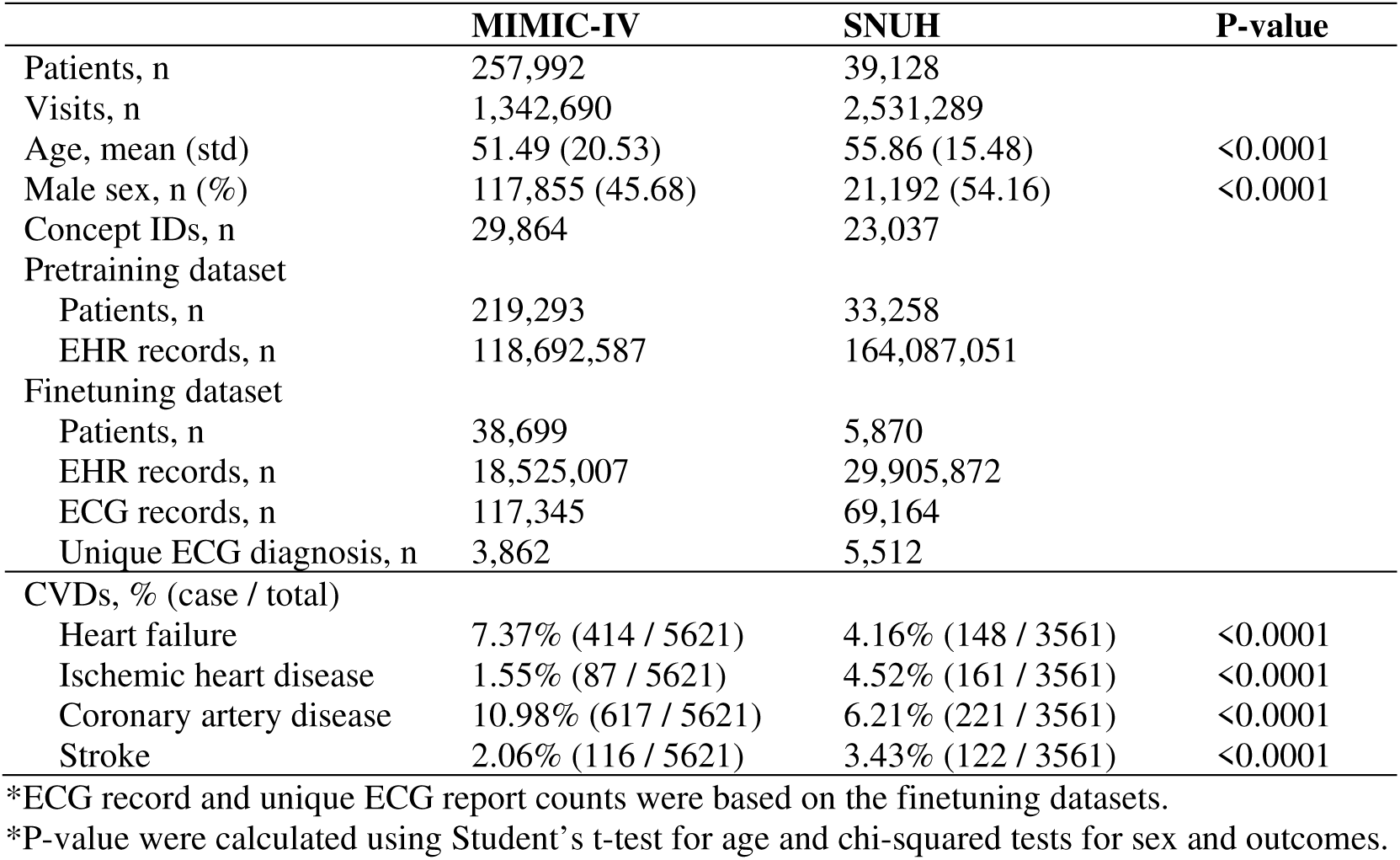
Baseline characteristics of finetuning datasets.

### TRIM representation analysis

We first examined how well ECG records from ECG and EHR systems were aligned (Fig. 2a). For MIMIC-IV, only 3,030 records overlapped, among 117,345 records from the ECG system and 4,071 records from the EHR system. For SNUH, 33,620 records were recorded in both ECG and EHR systems, while 35,544 records from the ECG system and 6,329 records from the EHR system did not overlap. These results indicate that ECG measurements are not routinely recorded in the EHR system, and the ECG system may also miss records. Therefore, integrating records from the ECG system into patient trajectories could enhance the validity of multimodal EHR FM research.

**Fig 2.**
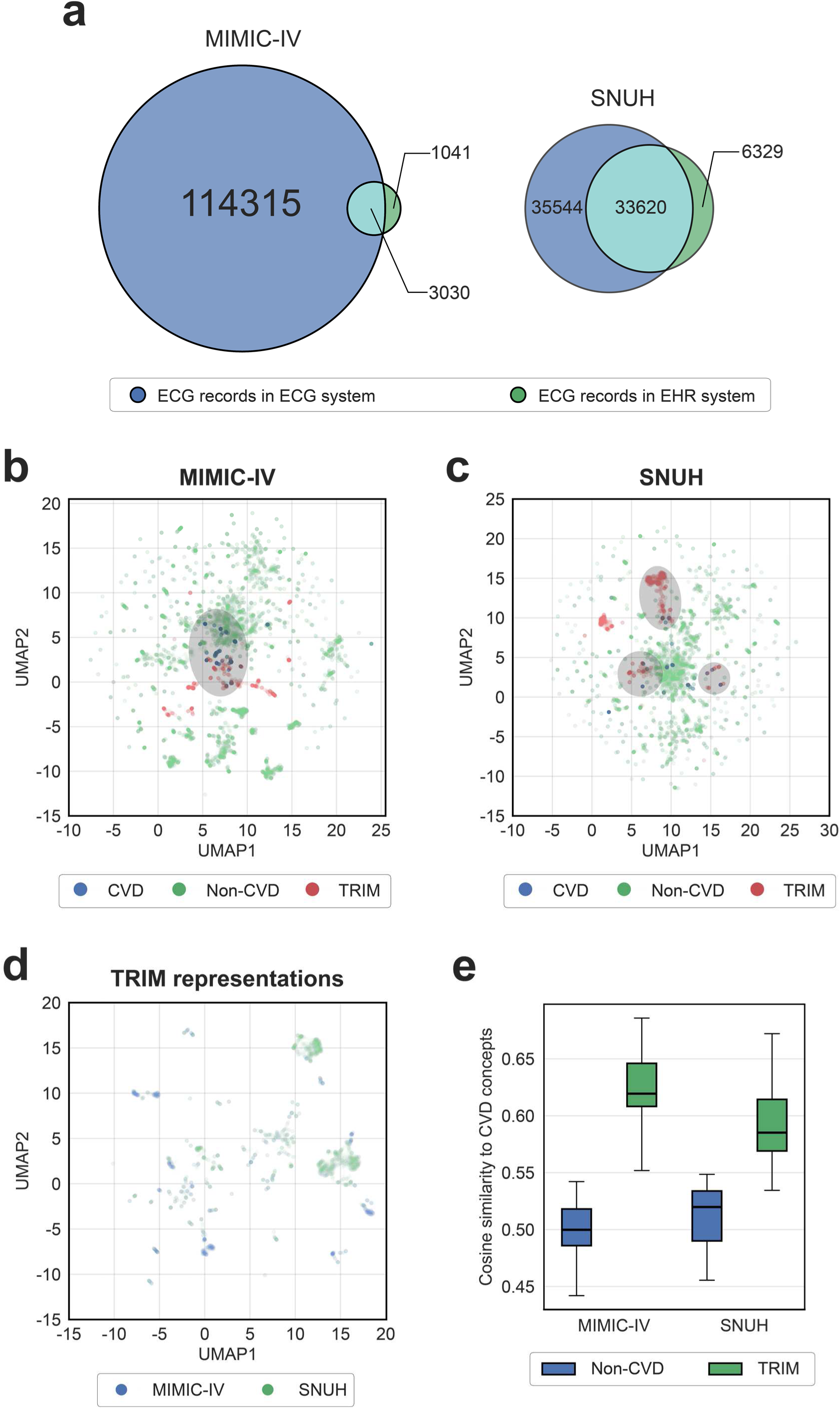
Overview of TRIM-style ECG representations. **a**, Venn diagram of ECG records in ECG and EHR systems. Larger overlapping area indicates better reflection of ECG records in the EHR system. **b**, UMAP visualization of CVD and non-CVD EHR concept representations and TRIM-style ECG representations of the MIMIC-IV dataset. To enhance visibility given the low proportion of CVD concepts, their corresponding points were displayed 50 times darker than other representations. **c**, UMAP visualization of CVD and non-CVD concept representations and TRIM-style ECG representations of the SNUH dataset. To enhance visibility given the low proportion of CVD concepts, their corresponding points were displayed 50 times darker than other representations. **d**, UMAP visualization of TRIM-style ECG representations of the MIMIC-IV and SNUH datasets. **e**, Boxplots of cosine similarity to CVD concepts for non-CVD EHR concept representations and TRIM-style ECG representations.

Next, we analyzed the characteristics of TRIM representations through UMAP visualization^13^. In MIMIC-IV, CVD concept representations formed a clear cluster, and TRIM representations were closely located within this cluster, as shown in the shaded region in Fig. 2b. However, TRIM representations were not fully covered by CVD concept representations, demonstrating their distinctiveness and diversity. In contrast, non-CVD concept representations were widely scattered without specific patterns.

In SNUH, CVD concept representations did not form a distinct cluster, but TRIM representations were closely located with them, as shown in the shaded regions in Fig. 2c. Compared to MIMIC-IV, SNUH TRIM representations formed clearer clusters, demonstrating less diversity. Non-CVD concept representations were also widely scattered without specific patterns, similar to MIMIC-IV. Since TRIM representations were generated from ECG diagnosis descriptions, they inherently contained cardiovascular information, explaining their proximity to CVD concept representations in both datasets.

TRIM representations from MIMIC-IV and SNUH formed separate, non-overlapping clusters (Fig. 2d), reflecting differences in the reporting practices of ECG systems. MIMIC-IV TRIM representations showed a scattered pattern with only several small clusters, while SNUH TRIM representations formed several large, distinct clusters. Despite thousands of unique ECG diagnoses in each dataset, only a few major clusters were observed, indicating common reporting patterns of cardiovascular conditions. Nevertheless, many irregularly scattered representations existed outside these clusters, reflecting diverse or rare ECG findings.

Quantitatively, cosine similarity to CVD concepts was also higher for TRIM representations than for non-CVD concept representations across both datasets, consistent with the UMAP visualization patterns (Fig. 2e).

### CVD prediction performance evaluation

We evaluated CVD prediction performance on seven models: (1) raw ECG model^14^, (2) EHR FM^15^, (3) EHR FM with raw ECG signals, (4) MedRep-based EHR FM, (5) MedRep-based EHR FM with raw ECG signals, (6) MedRep-based EHR FM with TRIM, and (7) MedRep-based EHR FM with TRIM and raw ECG signals (Table 2). For evaluation metrics, we used macro area under the receiver operating characteristic curve (AUROC), macro area under the precision-recall curve (AUPRC), and macro F1-score. The MedRep-based EHR FM with TRIM and raw ECG signals achieved the best performance across both datasets and all metrics with AUROC of 0.729, AUPRC of 0.159, and F1-score of 0.190 in MIMIC-IV, and AUROC of 0.764, AUPRC of 0.204, and F1-score of 0.216 in SNUH. The adoption of TRIM improved overall predictive performance for both models with and without raw ECG signals, across both datasets. MedRep representations substantially improved performance compared to the baseline EHR FM, and adding TRIM further enhanced predictive accuracy beyond MedRep alone, demonstrating the complementary value of structured ECG diagnostic information. Disease-specific prediction performance for each CVD is summarized in Supplementary Tables 1-4. The models with TRIM achieved the best performance across all metrics for all CVDs, except for the IHD cohort in MIMIC-IV.

**Table 2.** CVD prediction performance. Experiments were performed with five random seeds and all values are shown as mean ± SD.

| Model | Dataset 1: MIMIC-IV |  |  | Dataset 2: SNUH |  |  |
| --- | --- | --- | --- | --- | --- | --- |
|  | AUROC | AUPRC | F1 | AUROC | AUPRC | F1 |
| Raw ECG | 0.623 | 0.097 | 0.139 | 0.553 | 0.078 | 0.123 |
| | $\pm 0.085$ | $\pm 0.018$ | $\pm 0.027$ | $\pm 0.036$ | $\pm 0.019$ | $\pm 0.017$ |
| EHR FM | 0.611 | 0.108 | 0.165 | 0.680 | 0.130 | 0.179 |
| | $\pm 0.080$ | $\pm 0.017$ | $\pm 0.016$ | $\pm 0.033$ | $\pm 0.022$ | $\pm 0.021$ |
| EHR FM + Raw ECG | 0.656 | 0.113 | 0.168 | 0.683 | 0.134 | 0.183 |
| | $\pm 0.028$ | $\pm 0.007$ | $\pm 0.014$ | $\pm 0.013$ | $\pm 0.014$ | $\pm 0.018$ |
| EHR FM + MedRep | 0.692 | 0.140 | 0.182 | 0.724 | 0.159 | 0.193 |
| | $\pm 0.046$ | $\pm 0.020$ | $\pm 0.016$ | $\pm 0.044$ | $\pm 0.019$ | $\pm 0.017$ |
| EHR FM + MedRep + Raw ECG | 0.692 | 0.134 | 0.179 | 0.737 | 0.182 | 0.200 |
| | $\pm 0.022$ | $\pm 0.015$ | $\pm 0.015$ | $\pm 0.020$ | $\pm 0.029$ | $\pm 0.014$ |
| EHR FM + MedRep + TRIM | 0.711 | 0.150 | 0.186 | 0.742 | 0.198 | 0.209 |
| | $\pm 0.039$ | $\pm 0.016$ | $\pm 0.009$ | $\pm 0.013$ | $\pm 0.021$ | $\pm 0.017$ |
| EHR FM + MedRep + TRIM + Raw ECG | **0.729** | **0.159** | **0.190** | **0.764** | **0.204** | **0.216** |
|  | **$\pm 0.019$** | **$\pm 0.011$** | **$\pm 0.008$** | **$\pm 0.022$** | **$\pm 0.017$** | **$\pm 0.016$** |

\*Bold indicates the best.
\*AUROC, area under the receiver operating characteristic curve; AUPRC, area under the precision-recall curve

### Model interpretation of the MIMIC-IV dataset

In this section, we first explore which components of the model mainly affected model decisions. We conducted a cross-modal correlation analysis between trajectory and ECG representations—the output vectors from EHR FM and ECG encoder (Fig. 1d)—by calculating the Pearson correlation coefficient (PCC) between all pairs of their 768 latent dimensions (Fig. 3a)^9^. The EHR FMs with TRIM exhibited more significant correlations than those without TRIM, with 223,328 (15.20%) and 119,856 (8.16%) significant correlations among out of 1,472,640 total dimension pairs (5 seeds × 768(768-1)/2 pairs), respectively, at p<0.01. This increase in cross-modal connections demonstrates that TRIM integration encourages better semantic alignment between trajectory and ECG representations, which may contribute to the improved and stable performance observed consistently across CVDs and datasets^9^.

**Fig. 3.**
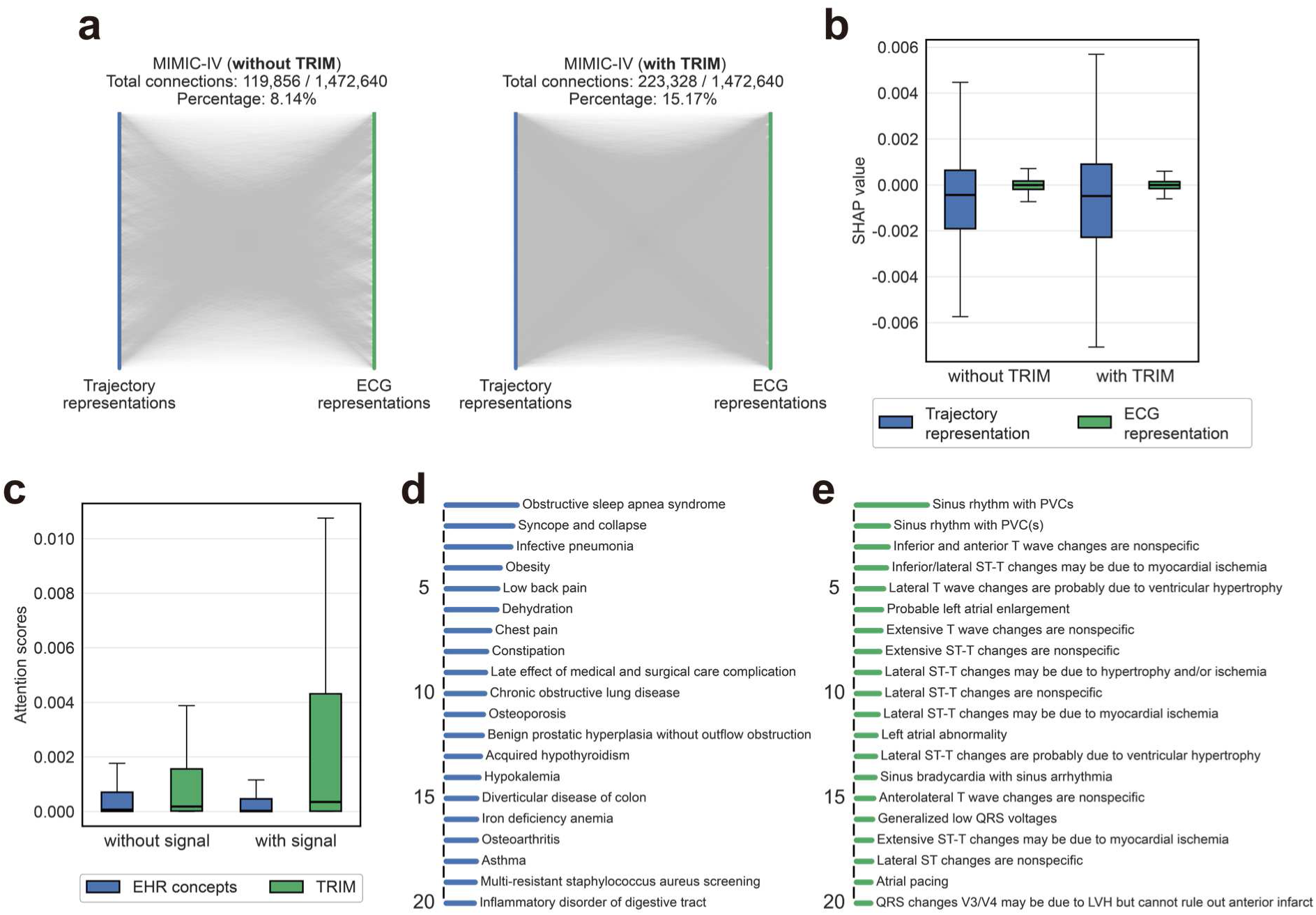
Model interpretation of the MIMIC-IV dataset. **a**, Cross-modal correlation analysis between trajectory and ECG representations. Lines represent statistically significant correlations (p<0.01). **b**, SHAP value distributions for trajectory and ECG representations. **c**, Attention score distributions for EHR concepts and TRIM within patient trajectories. **d**, Top 20 EHR concepts with the highest average attention scores. **e**, Top 20 TRIM records with the highest average attention scores.

To measure the impact of each latent representation on model decisions, we compared the SHAP values^16^ of trajectory and ECG representations (Fig. 3b). Both models with and without TRIM demonstrated that ECG representations had limited impact on predictions, with SHAP values concentrated near zero. In contrast, trajectory representation showed larger SHAP value distributions, indicating their dominant role in model decisions. Notably, the models with TRIM exhibited a wider SHAP value distribution for trajectory representations compared to those without TRIM, suggesting that TRIM integration amplified the influence of patient trajectory information on model predictions. While the combination of ECG and EHR improved overall performance, model decisions were predominantly driven by trajectory representations, as raw ECG signals alone were insufficient for accurate CVD prediction (Table 2).

In addition to latent space analysis, we explored attention scores within patient trajectories, as attention scores can reflect the importance of each record^15,17^. For both models with and without raw ECG signals, the attention scores for TRIM records were substantially higher than those for EHR concepts, indicating that ECG diagnosis descriptions integrated into patient trajectories played a more dominant role in the feature selection process of EHR FMs (Fig. 3c).

To examine which ECG diagnoses were influential in predicting CVDs, we reported the top 20 EHR concepts and TRIM records with the highest attention scores (Figs. 3d-3e). For EHR concepts, obstructive sleep apnea syndrome (OSA), which has close relationship with CVDs^18,19^, had the highest average attention score (Fig. 3d). Syncope and collapse^20,21^, obesity^22,23^, chronic obstructive lung disease (COPD)^24^, and acquired hypothyroidism^25^ have also been reported as risk factors for CVDs. Hypokalemia^26^ and iron deficiency anemia^27^ are risk factors of HF, and chest pain is a primary symptom of myocardial infarction (MI). Low back pain, osteoporosis, and benign prostatic hyperplasia without outflow obstruction are associated with older age and share common risk factors with CVDs; however, their direct relation to CVDs remains unclear.

Among TRIM records, premature ventricular complex (PVC), which is highly associated with HF and IHD^28,29^, had the highest attention score (Fig. 3e). Ischemia-related ST-T changes^30^, left atrial abnormality^31^, and ventricular hypertrophy-related changes^32^ are closely related to IHD, atrial fibrillation (AF), and HF, respectively. While the most frequent ECG diagnoses in the dataset were simple statements such as “Abnormal ECG”, “Sinus rhythm”, “Borderline ECG”, and “Normal ECG”, these diagnoses were not represented in the top 20 TRIM records. This result indicates that the model focused more on specific ECG conditions with clinical significance rather than generic diagnostic statements.

### Model interpretation of the SNUH dataset

Similar to MIMIC-IV, more significant correlations were observed in the EHR FMs with TRIM than those without TRIM, with 110,006 (7.47%) and 88,664 (6.02%) significant correlations, respectively (Fig. 4a). SHAP value distributions of trajectory and ECG representations also showed similar patterns to those of MIMIC-IV, although the distribution of trajectory representations in the models with TRIM was only slightly broader than those without TRIM compared to the MIMIC-IV result (Fig. 4b). The attention scores for TRIM records were also higher than those for EHR concepts across both models with and without raw ECG signals (Fig. 4c).

**Fig. 4.**
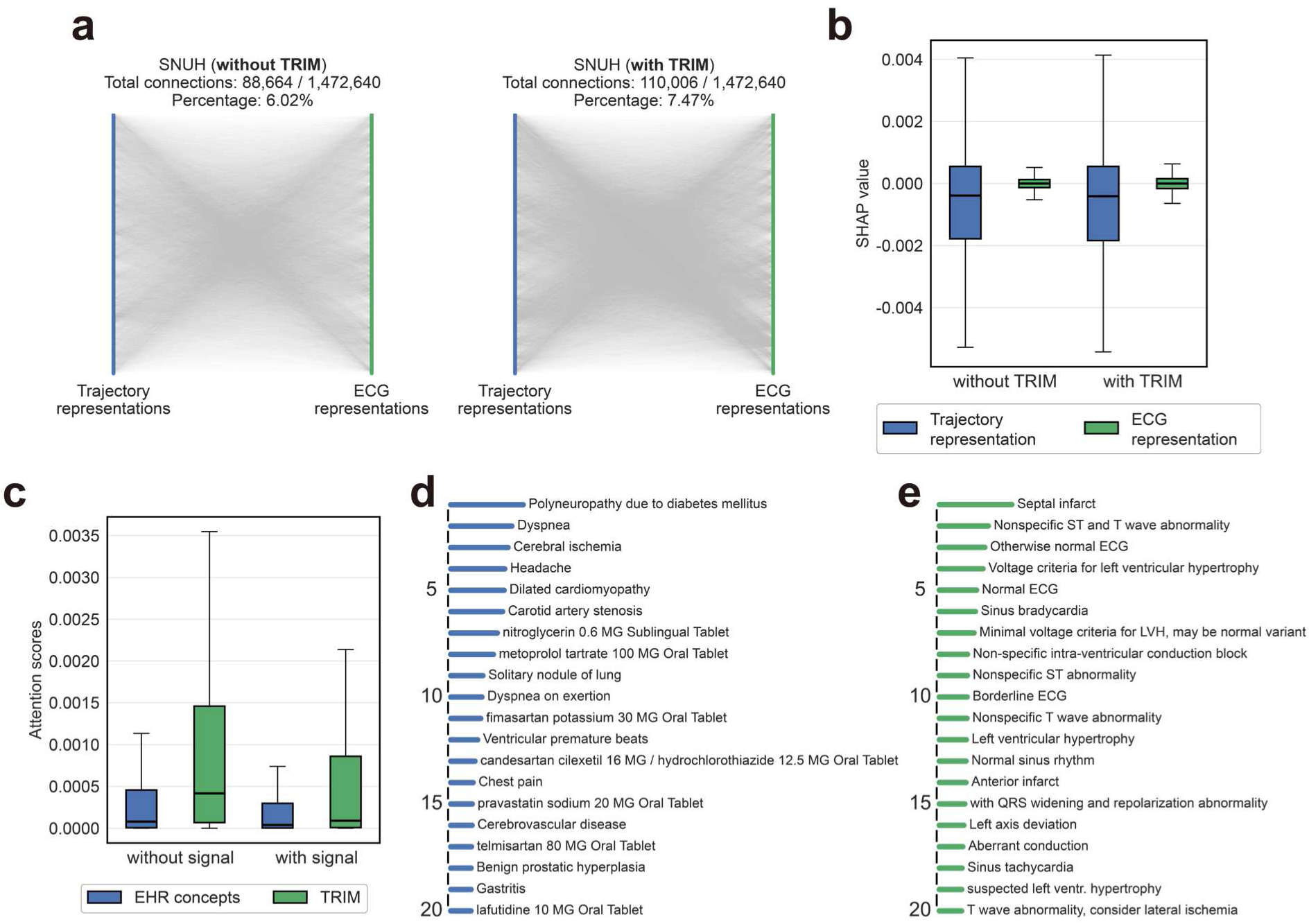
Model interpretation of the SNUH cohort. **a**, Cross-modal correlation analysis between trajectory and ECG representations. Lines represent statistically significant correlations (p<0.01). **b**, SHAP value distributions for trajectory and ECG representations. **c**, Attention score distributions for EHR concepts and TRIM within patient trajectories. **d**, Top 20 EHR concepts with the highest attention scores. **e**, Top 20 TRIM records with the highest attention scores.

For the top 20 EHR concepts, polyneuropathy due to diabetes mellitus, which is a major risk factor for CVD^33^, received the highest average attention score (Fig. 4d). Dyspnea is a cardinal symptom of HF and CAD^34^, while cerebral ischemia and carotid artery stenosis are primary risk factors for stroke. Several medications for CVD prevention and treatment were also included in the top 20. These drugs, including nitroglycerin^35^, metoprolol tartrate^36^, fimasartan potassium^37^, candesartan cilexetil^38^, pravastatin^39^, and telmisartan^40^, are used for CVD management, whereas lafutidine is a gastric acid suppressant. Benign prostatic hyperplasia and chest pain were common to both MIMIC-IV and SNUH. In contrast to MIMIC-IV, SNUH models focused more on stroke-related concepts and medication records regarding CVD management, while MIMIC-IV did not include any drug concept in the top 20.

Unlike the MIMIC-IV dataset, the top 20 TRIM records in SNUH included “Normal ECG” and “Normal sinus rhythm” (Fig. 4e). This was because 48.64% of MIMIC-IV trajectories contained these records, while only 1.52% of SNUH trajectories included them. Thus, the presence of “Normal ECG” and “Normal sinus rhythm” records represented distinctive features in SNUH, whereas MIMIC-IV models recognized them as common features. Similarly, “Borderline ECG” appeared in 53.18% of MIMIC-IV trajectories, but only in 17.69% of SNUH trajectories. Although “Abnormal ECG” was the most frequent ECG diagnosis in SNUH, it was not included in the top 20, similar to MIMIC-IV. This suggests that SNUH models also focused more on specific ECG conditions than generic diagnostic statements. Besides these generic records, several specific diagnoses, including septal infarct^30^, anterior infarct^41^, left ventricular hypertrophy (LVH)^32^, sinus bradycardia^42^, sinus tachycardia^43^, and left axis deviation^44^, were included, all of which are established indicators of CVD risk.

### Survival analysis for model decision validation

We conducted survival analysis to validate model decisions by fitting Cox proportional hazard (CPH) models, adjusted for age and sex, that regressed each CVD outcome (HF, IHD, CAD, and stroke) on model decisions (Figs. 5-6). Patients were classified into positive and negative groups based on thresholds determined by Youden’s index^45^, and Nelson-Aalen cumulative hazard curves were constructed for each CVD. Across all CVDs and models in both datasets, the gap between positive and negative groups increased proportionally over time, supporting the proportional hazards assumption. For models with raw ECG signals, TRIM integration generally widened the gap between risk groups (Figs. 5a and 6a). Models without raw ECG signals also exhibited a larger gap with TRIM, except for the IHD cohort in MIMIC-IV and the IHD and stroke cohorts in SNUH (Figs. 5c and 6c). In all cases, hazard ratios (HRs) of model decisions were statistically significant (Figs. 5b, 5d, 6b, and 6d). Notably, HRs consistently increased with TRIM integration, demonstrating stronger associations between positive predictions and actual CVD occurrence. All adjusted HRs for CVDs are summarized in Supplementary Tables 5-6.

**Fig. 5.**
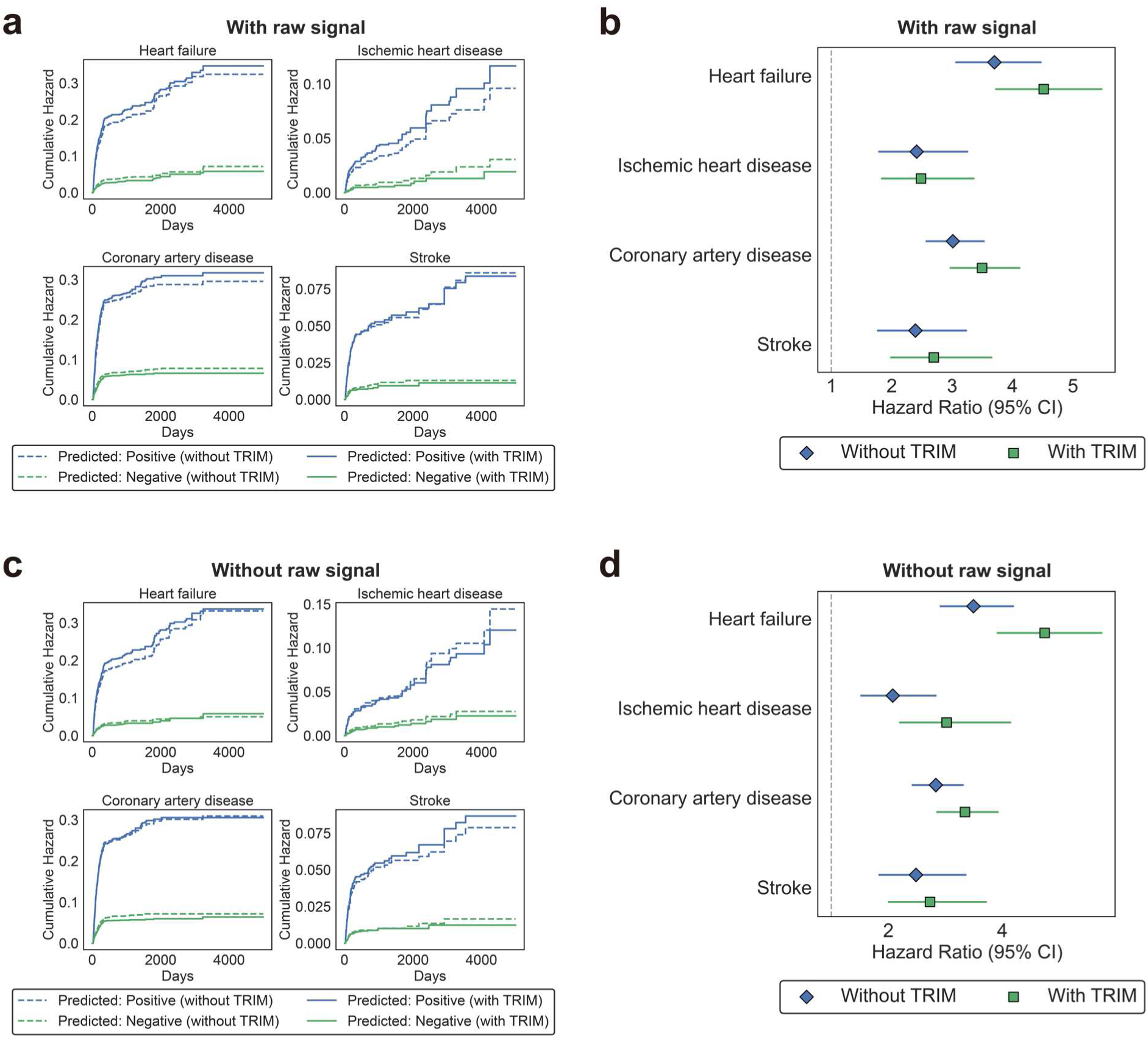
Survival analysis comparing models with and without TRIM on the MIMIC-IV dataset. **a**, Nelson-Aalen cumulative hazard curves for four CVDs in the models with raw ECG signals. **b**, Forest plot for HRs of the model decision with raw ECG signals, adjusted for age and sex. **c**, Nelson-Aalen cumulative hazard curves for four CVDs in the models without raw ECG signals. **d**, Forest plot for HRs of the model decision without raw ECG signals, adjusted for age and sex.

**Fig. 6.**
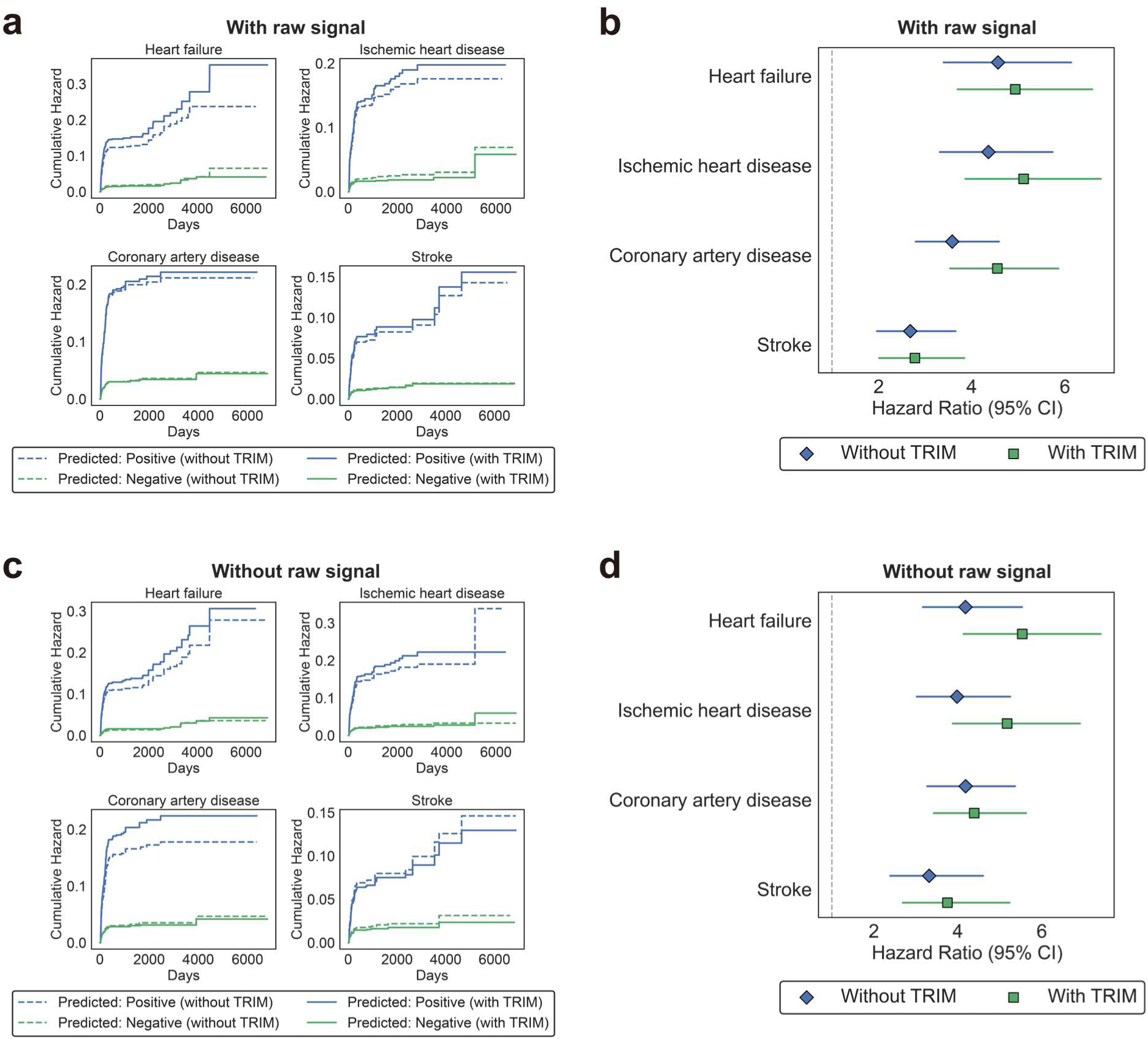
Survival analysis comparing models with and without TRIM on the SNUH dataset. **a**, Nelson-Aalen cumulative hazard curves for four CVDs in the models with raw ECG signals. **b**, Forest plot for HRs of the model decision with raw ECG signals, adjusted for age and sex. **c**, Nelson-Aalen cumulative hazard curves for four CVDs in the models without raw ECG signals. **d**, Forest plot for HRs of the model decision without raw ECG signals, adjusted for age and sex.

## Discussion

In this study, we proposed TRIM, a novel strategy for integrating unstructured medical data (e.g., ECG, CXR, CT) into EHR-based patient trajectories for multimodal EHR FMs. TRIM was motivated by two key observations: First, EHR FMs adopting text-based representations have achieved superior performance compared to traditional token-based models^11^. Second, unstructured medical data are typically accompanied by free-text diagnostic reports written by clinicians or automated systems. Leveraging this, we generated representations for unstructured data using their corresponding reports and MedRep’s text-based representation generator, thereby creating MedRep-style representations for ECG records that could be seamlessly integrated into patient trajectories.

TRIM addresses critical limitations of prior multimodal EHR FM frameworks. Previous studies (e.g., COMET^9^ and HAIM^10^) generated representations for each modality separately and integrated them only at the final prediction stage. This approach could not consider temporal relationships between different data types throughout the patient timeline. On the other hand, representing multiple unstructured data points may have computational challenges—in this study, each EHR record was a 768-dimensional vector, while a single ECG comprised 60,000-dimensional (12 lead × 500 Hz × 10 seconds). The lack of standardized methods for processing longitudinal unstructured data further limits the reliability of such approaches. Even when constructing unstructured data representations is possible, integrating them into EHR-based trajectories remains challenging due to semantic incompatibility across modalities. By converting unstructured data into compatible representations through MedRep’s text-based representation generator, TRIM representations aligned with related EHR concept representations (Fig. 2b-2c, 2e) and enabled EHR FMs to simultaneously analyze longitudinal structured and unstructured data while considering the time intervals between all records. We demonstrated the effectiveness of TRIM through comprehensive quantitative analyses (performance evaluation and survival analysis) and qualitative assessments (representation analysis and model interpretation).

While the models with TRIM and raw ECG signals achieved the best overall performance, the models with only TRIM showed the second-best performance with AUROC of 0.711, AUPRC of 0.150, F1-score of 0.186 in MIMIC-IV and AUROC of 0.742, AUPRC of 0.198, and F1-score of 0.209 in SNUH (Table 2). This result demonstrates that directly integrating longitudinal unstructured data into patient trajectories without requiring complex signal processing is more effective than integrating modality-specific representations at the final prediction stage (EHR FM with raw ECG signals and without TRIM). Furthermore, the combination of TRIM with raw ECG signals exhibited synergistic effects, suggesting that TRIM representations and signal features provide complementary information for CVD prediction.

The interpretability of TRIM-integrated models offers significant advantages for clinical practice. Our model identified clinically established risk factors for CVDs, demonstrating strong alignment with existing literature and clinical knowledge. This is essential for using artificial intelligence (AI) models in clinical practice because physicians are more inclined to trust and implement AI systems if they perceive the results as being more explainable or comprehensible^46^. Our approach, which integrates ECG records into patient trajectories, can provide clinicians with additional information to trace meaningful cardiovascular findings. The higher attention scores for TRIM records compared to EHR concepts (Figs. 3c and 4c) validate that the model properly selected cardiovascular-specific information when making decisions. Moreover, the cross-modal correlation analysis revealed that TRIM integration enhanced semantic alignment between trajectory and ECG representations (Figs. 3a and 4a), suggesting that our models made stable and enhanced decisions fully utilizing different data types.

The survival analysis validated the clinical utility of model predictions. Since we designed the model to predict CVDs within one year after the index date, the Nelson-Aalen curves exhibited steep slopes until around 365 days across all CVDs and models (Figs. 5a, 5c, 6a, 6c), yet the curves continued to show monotonically increasing gaps between CVD and non-CVD groups even after the monitoring period. This implies that long-term CVD risk can be assessed through our model and that patients currently without CVDs but predicted to develop them can be provided with proper lifestyle modifications and medical interventions to prevent the development of CVDs. Moreover, the higher HRs observed in the models with TRIM provide clinicians with greater confidence in implementing preventive interventions for high-risk patients identified by the model.

It is notable that TRIM can be applied to other modalities beyond ECG, since most medical images, including CXR, CT, and MRI, are accompanied by free-text radiology reports written by radiologists^47^. Additionally, there are many other unstructured medical data types accompanied by textual descriptions, including nursing notes, endoscopy reports, and echocardiogram reports^48^. In particular, the file size of medical images is usually much larger than ECG signals (e.g., a single chest CT scan can exceed 100 MB), making direct integration computationally challenging. However, the textual reports or metadata associated with these images are readily accessible. Although such metadata could be integrated into an EHR database, unstructured data and EHR records are often not compatible due to different management systems, as demonstrated in Fig. 2a. Therefore, TRIM provides an optimal solution for integrating longitudinal unstructured medical data with structured EHR records, enabling EHR FMs to achieve a more comprehensive understanding of patient trajectories.

Despite their cardiovascular relevance, the target CVDs (HF, IHD, CAD, and stroke) are not directly diagnosed by ECGs alone, and the definitive diagnostic process is associated with substantial costs: HF diagnosis requires comprehensive evaluation through echocardiography, the gold standard for assessing left ventricular function and structural abnormalities, with the median prices ranging from $204 to $2,588^49^. For IHD and CAD, diagnosis typically involves coronary angiography or CT angiography, with median costs ranging from $2,868 to $9,203 for coronary angiography^49^. Stroke diagnosis requires emergency brain imaging, predominantly through CT or MRI, with aggregate costs of post-emergency department diagnostic evaluation around $2,000^50^. As these processes entail significant economic burdens, reducing unnecessary testing and promoting the use of diagnostic evaluations in clearly indicated cases is essential. Since ECG testing costs are relatively lower than the above tests and range from $50 to $300 for standard 12-lead recordings in primary care settings, leveraging routinely collected ECG and EHR data for CVD risk stratification offers a cost-effective screening approach. Rather than requiring expensive diagnostic imaging as an initial assessment tool, our approach enables early risk identification using readily available longitudinal data, potentially reserving advanced imaging modalities for high-risk patients. This risk-stratified diagnostic paradigm could reduce healthcare expenditures, addressing the growing economic burden of cardiovascular imaging^50^.

This study has several limitations. First, we only used ECG for unstructured medical data. This choice was intentional to effectively evaluate TRIM by incorporating a target-related modality (ECG for CVD prediction). Future work should explore other modalities such as CXR and CT to further investigate more complex multimodal EHR FMs across diverse clinical outcomes. Second, while the original MedRep includes relational information between concepts, we only used a graph-free version containing textual information only, because integrating ECG records into the graph ontology of medical concepts was not feasible. Future research should investigate methods to fully utilize the relational information included in the original MedRep for enhanced representation learning. Third, we used a single ECG raw signal from the last measurement. As raw signals contain more detailed information than the metadata used for TRIM, strategies for merging longitudinal raw signal data and free-text diagnoses need to be explored. Fourth, we constructed TRIM at the diagnosis-level rather than the record-level. Most ECG records consisted of multiple diagnoses like “Normal ECG” and “Normal sinus rhythm”, or “Sinus rhythm” and “Left axis deviation”. This study treated these multiple diagnoses as attached but separate tokens, but record-level TRIM may capture more detailed diagnostic patterns.

## Methods

### Data curation

This study used data from 257,992 and 39,128 adult patients aged over 18 years from MIMIC-IV and SNUH, respectively. For SNUH, we randomly sampled 39,128 patients with at least one ECG record measured in the inpatient ECG room or emergency room to exclude healthy individuals who routinely undergo ECG testing for biennial health checkups in Korea^51^. The data period spanned from 15 October 2004 to 2 February 2024. The MIMIC-IV data period was between 2008 and 2022^12^. Both MIMIC-IV and SNUH datasets were based on the Observational Medical Outcomes Partnership (OMOP) Common Data Model (CDM) format. Since MedRep is based on OMOP format and MIMIC-IV is not originally formatted in OMOP CDM, we converted the original MIMIC-IV into OMOP format using the official OHDSI GitHub repository (https://github.com/OHDSI/MIMIC). We used data from 85% of patients for pretraining (70% for training and 15% for validation) and the remaining 15% was used for finetuning with a 6:2:2 ratio for training, validation, and hold-out test sets.

Among patients in the finetuning dataset, those without ECG were excluded from MIMIC-IV (SNUH patients were initially selected based on ECG availability, so none were excluded). We then extracted all available past ECG records of these patients, resulting in 117,345 and 69,164 records from MIMIC-IV and SNUH, respectively. We divided patients with ECG records according to CVD record existence. For patients with CVD records, the index date was defined as the date of the last ECG measurement before the first CVD occurrence. Patients whose first CVD did not occur between one month and one year after the index date were excluded (one-month washout period and one-year monitoring period). For patients without CVD records, the index date was defined as the date of the last ECG measurement. To ensure no CVD occurrence within the monitoring period, patients without any visits at least one year after the index date were excluded. For both CVD and non-CVD groups, patients with fewer than 30 EHR records were excluded. Finally, MIMIC-IV contained 1,005 CVD patients and 4,616 non-CVD patients, and SNUH contained 477 and 3,084 patients, respectively. The population flowcharts are shown in Supplementary Figure 1.

The Institutional Review Board (IRB) of Seoul National University Hospital (IRB approval No. 2409-073-1570) approved the study with a waiver of informed consent, considering that our study used retrospective and observational EHR data. The approval aligns with the principles outlined in the Declaration of Helsinki, the Korean Bioethics and Safety Act (Law No. 16372), and the Human Research Protection Program–Standard Operating Procedure of Seoul National University Hospital.

### Data preprocessing and outcomes

Each patient trajectory consisted of integer sequences of medical concept indices, age indices, visit indices, record indices, and domain indices, all sorted in order of time. The maximum trajectory length was set to 2048. Every trajectory started with a [CLS] token, and [SEP] tokens were not used as visit indices already distinguished different visits. Unique ECG diagnoses were added to the original vocabularies of MIMIC-IV and SNUH, and these ECG diagnoses were treated as medical concepts and merged into patient trajectories as shown in Fig. 1d. The vocabulary size was 33,726 (29,864 EHR concepts and 3,862 ECG diagnoses) for MIMIC-IV and 26,704 (21,192 EHR concepts and 5,512 ECG diagnoses) for SNUH.

For raw ECG signals, we followed the preprocessing process described in a previous study that used a one-dimensional ResNet-based model for ECG analysis^14^. Specifically, we first applied a fifth-order Butterworth filter with a frequency range between 0.05 and 150 Hz to remove noise and then scaled the filtered signal to range between -1 and 1 for normalization. Missing values were imputed with 0. The dimension of ECG signals in both MIMIC-IV and SNUH was 12 × 5000.

For CVD outcomes, we used the ICD-10 taxonomy and converted the codes into OMOP concepts: I50 for HF; I20, I21, and I24 for IHD; I25 for CAD; and I60, I61, and I63 for stroke. OMOP concepts for each CVD are summarized in Supplementary Table 7.

### Generating descriptions for ECG diagnoses

TRIM representations were generated using LLM-enriched ECG diagnosis descriptions and the text-based representation generator of MedRep. ECG diagnoses are phrase-level, for example, “RBBB with left anterior fascicular block”. These phrase-level diagnoses were enriched through LLMs as shown in Figs. 1b-1c. Instead of training a new representation generating model, we used the text-based representation generator of MedRep which was trained on descriptions of more than 7 million medical concepts^11^ to align new ECG representations with original concept representations. The prompt for ECG diagnosis description was as follows: “Instruction: Briefly explain the clinical background and regarding treatments of each ECG diagnostic report with less than 5 sentences. Do not include sentences that are too ordinary (such as “further details would depend on the specific situation”) and focus on describing the representative clinical features of the concept. If possible, avoid using negation.” Generating descriptions for 9,374 (3,862 from MIMIC-IV and 5,512 from SNUH) unique diagnoses costed $0.95 with ChatGPT-4o-mini (OpenAI Inc). Examples of ECG diagnosis descriptions are provided in Supplementary Table 8.

### Model and training details

The architecture of the EHR FMs was based on CDM-BERT^15^, which introduces domain embedding to indicate the domain of masked tokens during pretraining, thereby preventing the model from finding unnecessary medical concepts from other domains. We used a 768-dimensional hidden dimension consistent with MedRep, 12 attention layers, and 12 attention heads. For the ECG encoder, we used a one-dimensional ResNet-based model. The detailed configuration of the ECG encoder is summarized in Supplementary Table 9.

For pretraining, we selected the models with the lowest masked language model (MLM) loss during 50-epoch training. The pretraining configuration was as follows: a batch size of 32, a learning rate of 0.00005, and an AdamW optimizer with a weight decay of 0.01. Patient trajectories exceeding the maximum length were sliced into non-overlapping sub-trajectories to avoid potential dependency among trajectories from a single patient. Pretraining was conducted using 8 NVIDIA RTX A6000 GPUs, requiring approximately 18 hours for MIMIC-IV and 12 hours for SNUH. Although SNUH had substantially fewer patients for pretraining than MIMIC-IV (33,258 vs. 219,293), it contained considerably more EHR records per patient (4,934 vs. 541), resulting in many sliced sub-trajectories. Consequently, the training time for SNUH was approximately 67% that of MIMIC-IV.

For finetuning, we trained each model for 50 epochs with an early stopping patience of 10 to prevent overfitting. The loss function was binary cross entropy for each target, and early stopping was applied based on the macro AUROC. We conducted finetuning using five random seeds, randomly splitting the data into training, validation, and hold-out test sets. All performance evaluation processes were performed on the hold-out test sets. Each finetuning task was performed with a single NVIDIA RTX A6000 GPU, requiring 1-4 hours per random seed.

### Model interpretation

To explore which components of the model mainly affected model decisions, we conducted a two-fold analysis: latent space analysis and attention score analysis. For latent space analysis, we conducted a cross-modal correlation analysis between trajectory and ECG representations by calculating the PCC between all pairs of their 768 latent dimensions, to evaluate the alignment between these representations. In addition, we calculated SHAP values^16^ of trajectory and ECG representations to compare the impact of each latent representation on model decisions. We used an Integrated Gradients-based SHAP implementation^52^ (GradientExplainer in the SHAP package) with a background sample size of 100.

For attention score analysis, we used the attention vector of each [CLS] token from the last attention layer output and averaged the attention scores for each concept. For qualitative analysis of concepts with high average attention scores, we only included concepts that appeared in more than 100 patients, to avoid rare concepts coincidentally having extremely high attention scores.

All model interpretation processes were performed on the hold-out test sets. As we conducted experiments with five random seeds, we reported the results of both latent space analysis and attention score analysis after merging the five results.

### Statistical analysis

Age between the datasets were compared by calculating p-values using the Student’s t-test and sex and CVD incidence ratio between the datasets were compared by chi-squared test. For performance metrics, we used the AUROC, AUPRC, and F1-score, and the confidence intervals (CIs) of the AUROC and AUPRC were calculated using DeLong’s method, and the CIs for F1-score was calculated using Wilson’s method. For survival analysis, CPH models were fitted by Breslow’s method with a penalizer of 0.01. Statistical significance was set at α = 0.05. All statistical analyses were performed using Python (version 3.9.19), and Lifelines (version 0.30.0) in Python was used for survival analysis.

## Data Availability

MIMIC-IV is publicly available upon application for access at https://physionet.org/content/mimiciv/2.2/. The SNUH dataset is not publicly available due to patient privacy and is available from the corresponding author on reasonable request and IRB approvals. The MedRep dataset is available in our GitHub repository at https://github.com/kicarussays/MedRep.

## Code Availability

The source code used in this study is publicly available at https://github.com/kicarussays/TRIM.

## Supplementary Materials

**Supplementary Table 1.** HF prediction performance. Experiments were performed with five random seeds and all values are shown as mean ± SD.

| Model | Dataset 1: MIMIC-IV |  |  | Dataset 2: SNUH |  |  |
| --- | --- | --- | --- | --- | --- | --- |
|  | AUROC | AUPRC | F1 | AUROC | AUPRC | F1 |
| Raw ECG | 0.662 | 0.160 | 0.213 | 0.595 | 0.124 | 0.135 |
| | $\pm 0.156$ | $\pm 0.044$ | $\pm 0.076$ | $\pm 0.082$ | $\pm 0.063$ | $\pm 0.038$ |
| EHR FM | 0.635 | 0.163 | 0.247 | 0.696 | 0.137 | 0.168 |
| | $\pm 0.055$ | $\pm 0.024$ | $\pm 0.024$ | $\pm 0.026$ | $\pm 0.038$ | $\pm 0.027$ |
| EHR FM + Raw ECG | 0.705 | 0.183 | 0.258 | 0.721 | 0.126 | 0.172 |
| | $\pm 0.047$ | $\pm 0.035$ | $\pm 0.027$ | $\pm 0.019$ | $\pm 0.023$ | $\pm 0.027$ |
| EHR FM + MedRep | 0.725 | 0.203 | 0.254 | 0.774 | 0.163 | 0.184 |
| | $\pm 0.059$ | $\pm 0.041$ | $\pm 0.029$ | $\pm 0.049$ | $\pm 0.041$ | $\pm 0.028$ |
| EHR FM + MedRep + Raw ECG | 0.740 | 0.230 | 0.265 | 0.760 | 0.156 | 0.203 |
| | $\pm 0.025$ | $\pm 0.035$ | $\pm 0.036$ | $\pm 0.024$ | $\pm 0.026$ | $\pm 0.018$ |
| EHR FM + MedRep + TRIM | 0.766 | 0.250 | 0.279 | 0.783 | 0.191 | 0.205 |
| | $\pm 0.046$ | $\pm 0.039$ | $\pm 0.031$ | $\pm 0.043$ | $\pm 0.076$ | $\pm 0.033$ |
| EHR FM + MedRep + TRIM + Raw ECG | **0.785** | **0.278** | **0.296** | **0.787** | **0.208** | **0.230** |
|  | **$\pm 0.026$** | **$\pm 0.029$** | **$\pm 0.026$** | **$\pm 0.048$** | **$\pm 0.044$** | **$\pm 0.026$** |

\*Bold indicates the best.
\*AUROC, area under the receiver operating characteristic curve; AUPRC, area under the precision-recall curve

**Supplementary Table 2.** IHD prediction performance. Experiments were performed with five random seeds and all values are shown as mean ± SD.

| Model | Dataset 1: MIMIC-IV |  |  | Dataset 2: SNUH |  |  |
| --- | --- | --- | --- | --- | --- | --- |
|  | AUROC | AUPRC | F1 | AUROC | AUPRC | F1 |
| Raw ECG | 0.604 | 0.033 | 0.046 | 0.544 | 0.062 | 0.117 |
| | $\pm 0.091$ | $\pm 0.018$ | $\pm 0.008$ | $\pm 0.021$ | $\pm 0.005$ | $\pm 0.016$ |
| EHR FM | 0.567 | 0.034 | **0.078** | 0.657 | 0.147 | 0.191 |
| | $\pm 0.108$ | $\pm 0.010$ | **$\pm 0.033$** | $\pm 0.078$ | $\pm 0.047$ | $\pm 0.047$ |
| EHR FM + Raw ECG | 0.608 | 0.043 | 0.061 | 0.689 | 0.161 | 0.202 |
| | $\pm 0.062$ | $\pm 0.023$ | $\pm 0.017$ | $\pm 0.051$ | $\pm 0.036$ | $\pm 0.034$ |
| EHR FM + MedRep | 0.635 | **0.058** | 0.067 | 0.737 | 0.196 | 0.222 |
| | $\pm 0.077$ | **$\pm 0.030$** | $\pm 0.023$ | $\pm 0.072$ | $\pm 0.031$ | $\pm 0.026$ |
| EHR FM + MedRep + Raw ECG | 0.643 | 0.033 | 0.050 | 0.745 | 0.222 | 0.208 |
| | $\pm 0.068$ | $\pm 0.012$ | $\pm 0.021$ | $\pm 0.065$ | $\pm 0.047$ | $\pm 0.023$ |
| EHR FM + MedRep + TRIM | 0.655 | 0.032 | 0.053 | 0.760 | 0.233 | **0.242** |
| | $\pm 0.059$ | $\pm 0.011$ | $\pm 0.013$ | $\pm 0.039$ | $\pm 0.063$ | **$\pm 0.040$** |
| EHR FM + MedRep + TRIM + Raw ECG | **0.683** | 0.050 | 0.054 | **0.797** | **0.245** | 0.231 |
| | **$\pm 0.045$** | $\pm 0.021$ | $\pm 0.011$ | **$\pm 0.042$** | **$\pm 0.062$** | $\pm 0.053$ |

\*Bold indicates the best.
\*AUROC, area under the receiver operating characteristic curve; AUPRC, area under the precision-recall curve

**Supplementary Table 3.** CAD prediction performance. Experiments were performed with five random seeds and all values are shown as mean ± SD.

| Model | Dataset 1: MIMIC-IV |  |  | Dataset 2: SNUH |  |  |
| --- | --- | --- | --- | --- | --- | --- |
|  | AUROC | AUPRC | F1 | AUROC | AUPRC | F1 |
| Raw ECG | 0.595 | 0.162 | 0.237 | 0.552 | 0.082 | 0.150 |
| | $\pm 0.067$ | $\pm 0.022$ | $\pm 0.033$ | $\pm 0.034$ | $\pm 0.008$ | $\pm 0.017$ |
| EHR FM | 0.638 | 0.203 | 0.264 | 0.688 | 0.163 | 0.242 |
| | $\pm 0.110$ | $\pm 0.051$ | $\pm 0.068$ | $\pm 0.047$ | $\pm 0.038$ | $\pm 0.051$ |
| EHR FM + Raw ECG | 0.641 | 0.178 | 0.281 | 0.685 | 0.179 | 0.251 |
| | $\pm 0.030$ | $\pm 0.018$ | $\pm 0.025$ | $\pm 0.028$ | $\pm 0.026$ | $\pm 0.052$ |
| EHR FM + MedRep | 0.701 | 0.245 | 0.326 | 0.717 | 0.190 | 0.233 |
| | $\pm 0.035$ | $\pm 0.030$ | $\pm 0.031$ | $\pm 0.077$ | $\pm 0.036$ | $\pm 0.039$ |
| EHR FM + MedRep + Raw ECG | 0.681 | 0.234 | 0.315 | 0.725 | 0.253 | 0.264 |
| | $\pm 0.059$ | $\pm 0.038$ | $\pm 0.010$ | $\pm 0.057$ | $\pm 0.061$ | $\pm 0.036$ |
| EHR FM + MedRep + TRIM | 0.711 | **0.270** | **0.327** | **0.749** | **0.274** | **0.270** |
| | $\pm 0.038$ | $\pm 0.021$ | $\pm 0.020$ | $\pm 0.039$ | $\pm 0.044$ | $\pm 0.029$ |
| EHR FM + MedRep + TRIM + Raw ECG | **0.726** | 0.252 | **0.327** | 0.737 | 0.248 | 0.267 |
| | **$\pm 0.014$** | $\pm 0.026$ | **$\pm 0.015$** | $\pm 0.049$ | $\pm 0.036$ | $\pm 0.036$ |

\*Bold indicates the best.
\*AUROC, area under the receiver operating characteristic curve; AUPRC, area under the precision-recall curve

**Supplementary Table 4.** Stroke prediction performance. Experiments were performed with five random seeds and all values are shown as mean ± SD.

| Model | Dataset 1: MIMIC-IV |  |  | Dataset 2: SNUH |  |  |
| --- | --- | --- | --- | --- | --- | --- |
|  | AUROC | AUPRC | F1 | AUROC | AUPRC | F1 |
| Raw ECG | 0.631 | 0.033 | 0.061 | 0.523 | 0.046 | 0.088 |
| | $\pm 0.047$ | $\pm 0.002$ | $\pm 0.008$ | $\pm 0.067$ | $\pm 0.016$ | $\pm 0.017$ |
| EHR FM | 0.604 | 0.034 | 0.070 | 0.680 | 0.070 | 0.114 |
| | $\pm 0.086$ | $\pm 0.012$ | $\pm 0.011$ | $\pm 0.028$ | $\pm 0.014$ | $\pm 0.018$ |
| EHR FM + Raw ECG | 0.670 | 0.048 | 0.071 | 0.635 | 0.070 | 0.106 |
| | $\pm 0.038$ | $\pm 0.016$ | $\pm 0.014$ | $\pm 0.031$ | $\pm 0.013$ | $\pm 0.016$ |
| EHR FM + MedRep | 0.705 | 0.052 | 0.081 | 0.668 | 0.088 | 0.133 |
| | $\pm 0.065$ | $\pm 0.019$ | $\pm 0.023$ | $\pm 0.041$ | $\pm 0.029$ | $\pm 0.034$ |
| EHR FM + MedRep + Raw ECG | 0.704 | 0.041 | **0.085** | 0.719 | 0.098 | 0.124 |
| | $\pm 0.037$ | $\pm 0.010$ | **$\pm 0.019$** | $\pm 0.028$ | $\pm 0.020$ | $\pm 0.010$ |
| EHR FM + MedRep + TRIM | 0.712 | 0.048 | 0.084 | 0.677 | 0.096 | 0.118 |
| | $\pm 0.039$ | $\pm 0.008$ | $\pm 0.015$ | $\pm 0.040$ | $\pm 0.012$ | $\pm 0.027$ |
| EHR FM + MedRep + TRIM + Raw ECG | **0.724** | **0.056** | 0.081 | **0.734** | **0.114** | **0.136** |
| | **$\pm 0.054$** | **$\pm 0.028$** | $\pm 0.014$ | **$\pm 0.038$** | **$\pm 0.030$** | **$\pm 0.015$** |

\*Bold indicates the best.
\*AUROC, area under the receiver operating characteristic curve; AUPRC, area under the precision-recall curve

**Supplementary Table 5.** Adjusted HRs for CVDs (MIMIC-IV).

| Dataset: MIMIC-IV |  |  |  |  |  |  |  |  |  |  |
| --- | --- | --- | --- | --- | --- | --- | --- | --- | --- | --- |
| TRIM | ECG Signals | Variable | Outcome: HF<br>Hazard Ratio<br>(CI 95%) | p | Outcome: IHD<br>Hazard Ratio<br>(CI 95%) | p | Outcome: CAD<br>Hazard Ratio<br>(CI 95%) | p | Outcome: Stroke<br>Hazard Ratio<br>(CI 95%) | p |
| X | X | Age | 1.018<br>(1.013-1.024) | 0.0000 | 1.015<br>(1.007-1.023) | 0.0004 | 1.018<br>(1.013-1.022) | 0.0000 | 1.030<br>(1.021-1.039) | 0.0000 |
|  |  | Sex | 0.995<br>(0.845-1.171) | 0.9488 | 1.013<br>(0.762-1.348) | 0.9274 | 0.965<br>(0.833-1.117) | 0.6317 | 0.953<br>(0.720-1.262) | 0.7390 |
|  |  | Model decision | 3.701<br>(3.066-4.467) | 0.0000 | 2.412<br>(1.789-3.252) | 0.0000 | 3.012<br>(2.573-3.526) | 0.0000 | 2.391<br>(1.769-3.233) | 0.0000 |
| ✓ | X | Age | 1.016<br>(1.011-1.021) | 0.0000 | 1.014<br>(1.005-1.022) | 0.0011 | 1.016<br>(1.012-1.021) | 0.0000 | 1.029<br>(1.020-1.038) | 0.0000 |
|  |  | Sex | 0.993<br>(0.844-1.169) | 0.9308 | 1.017<br>(0.764-1.353) | 0.9087 | 0.967<br>(0.834-1.119) | 0.6495 | 0.962<br>(0.727-1.273) | 0.7857 |
|  |  | Model decision | 4.516<br>(3.727-5.473) | 0.0000 | 2.483<br>(1.835-3.359) | 0.0000 | 3.496<br>(2.971-4.115) | 0.0000 | 2.695<br>(1.989-3.652) | 0.0000 |
| X | ✓ | Age | 1.019<br>(1.014-1.024) | 0.0000 | 1.015<br>(1.007-1.024) | 0.0003 | 1.019<br>(1.014-1.024) | 0.0000 | 1.030<br>(1.021-1.039) | 0.0000 |
|  |  | Sex | 1.023<br>(0.869-1.204) | 0.7820 | 1.030<br>(0.774-1.370) | 0.8419 | 0.961<br>(0.830-1.113) | 0.5969 | 0.965<br>(0.729-1.277) | 0.8007 |
|  |  | Model decision | 3.493<br>(2.913-4.188) | 0.0000 | 2.076<br>(1.523-2.829) | 0.0000 | 2.832<br>(2.423-3.309) | 0.0000 | 2.484<br>(1.839-3.355) | 0.0000 |
| ✓ | ✓ | Age | 1.016<br>(1.011-1.021) | 0.0000 | 1.012<br>(1.004-1.021) | 0.0037 | 1.017<br>(1.013-1.022) | 0.0000 | 1.029<br>(1.020-1.037) | 0.0000 |
|  |  | Sex | 1.014<br>(0.862-1.194) | 0.8659 | 1.027<br>(0.771-1.366) | 0.8575 | 0.963<br>(0.832-1.116) | 0.6184 | 0.949<br>(0.717-1.256) | 0.7157 |
|  |  | Model decision | 4.741<br>(3.916-5.738) | 0.0000 | 3.020<br>(2.202-4.141) | 0.0000 | 3.344<br>(2.852-3.921) | 0.0000 | 2.730<br>(2.006-3.715) | 0.0000 |

**Supplementary Table 6.** Adjusted HRs for CVDs (SNUH).

| Dataset: SNUH |  |  |  |  |  |  |  |  |  |  |
| --- | --- | --- | --- | --- | --- | --- | --- | --- | --- | --- |
| TRIM | ECG Signals | Variable | Outcome: HF<br>Hazard Ratio<br>(CI 95%) | p | Outcome: IHD<br>Hazard Ratio<br>(CI 95%) | p | Outcome: CAD<br>Hazard Ratio<br>(CI 95%) | p | Outcome: Stroke<br>Hazard Ratio<br>(CI 95%) | p |
| X | X | Age | 1.030<br>(1.020-1.040) | 0.0000 | 1.028<br>(1.018-1.038) | 0.0000 | 1.028<br>(1.019-1.036) | 0.0000 | 1.022<br>(1.011-1.033) | 0.0000 |
|  |  | Sex | 1.008<br>(0.778-1.308) | 0.9491 | 0.814<br>(0.629-1.055) | 0.1200 | 0.768<br>(0.607-0.973) | 0.0285 | 1.161<br>(0.862-1.565) | 0.3258 |
|  |  | Model decision | 4.563<br>(3.395-6.133) | 0.0000 | 4.359<br>(3.313-5.735) | 0.0000 | 3.580<br>(2.796-4.583) | 0.0000 | 2.678<br>(1.963-3.654) | 0.0000 |
| ✓ | X | Age | 1.028<br>(1.018-1.038) | 0.0000 | 1.025<br>(1.015-1.035) | 0.0000 | 1.023<br>(1.014-1.032) | 0.0000 | 1.021<br>(1.010-1.032) | 0.0002 |
|  |  | Sex | 1.028<br>(0.793-1.333) | 0.8358 | 0.810<br>(0.625-1.049) | 0.1108 | 0.758<br>(0.598-0.959) | 0.0211 | 1.177<br>(0.873-1.586) | 0.2843 |
|  |  | Model decision | 4.934<br>(3.697-6.584) | 0.0000 | 5.113<br>(3.864-6.766) | 0.0000 | 4.550<br>(3.534-5.859) | 0.0000 | 2.778<br>(2.010-3.839) | 0.0000 |
| X | ✓ | Age | 1.032<br>(1.022-1.042) | 0.0000 | 1.028<br>(1.018-1.038) | 0.0000 | 1.025<br>(1.016-1.034) | 0.0000 | 1.019<br>(1.009-1.030) | 0.0004 |
|  |  | Sex | 1.042<br>(0.803-1.351) | 0.7581 | 0.804<br>(0.621-1.042) | 0.0989 | 0.749<br>(0.592-0.948) | 0.0164 | 1.167<br>(0.866-1.573) | 0.3107 |
|  |  | Model decision | 4.184<br>(3.165-5.530) | 0.0000 | 3.981<br>(3.020-5.248) | 0.0000 | 4.187<br>(3.268-5.363) | 0.0000 | 3.315<br>(2.388-4.603) | 0.0000 |
| ✓ | ✓ | Age | 1.026<br>(1.015-1.036) | 0.0000 | 1.025<br>(1.015-1.035) | 0.0000 | 1.025<br>(1.016-1.034) | 0.0000 | 1.017<br>(1.006-1.028) | 0.0027 |
|  |  | Sex | 1.043<br>(0.805-1.353) | 0.7491 | 0.822<br>(0.635-1.065) | 0.1389 | 0.757<br>(0.598-0.958) | 0.0204 | 1.170<br>(0.868-1.577) | 0.3020 |
|  |  | Model decision | 5.535<br>(4.140-7.400) | 0.0000 | 5.174<br>(3.875-6.909) | 0.0000 | 4.391<br>(3.426-5.627) | 0.0000 | 3.750<br>(2.688-5.232) | 0.0000 |

**Supplementary Figure 1.**
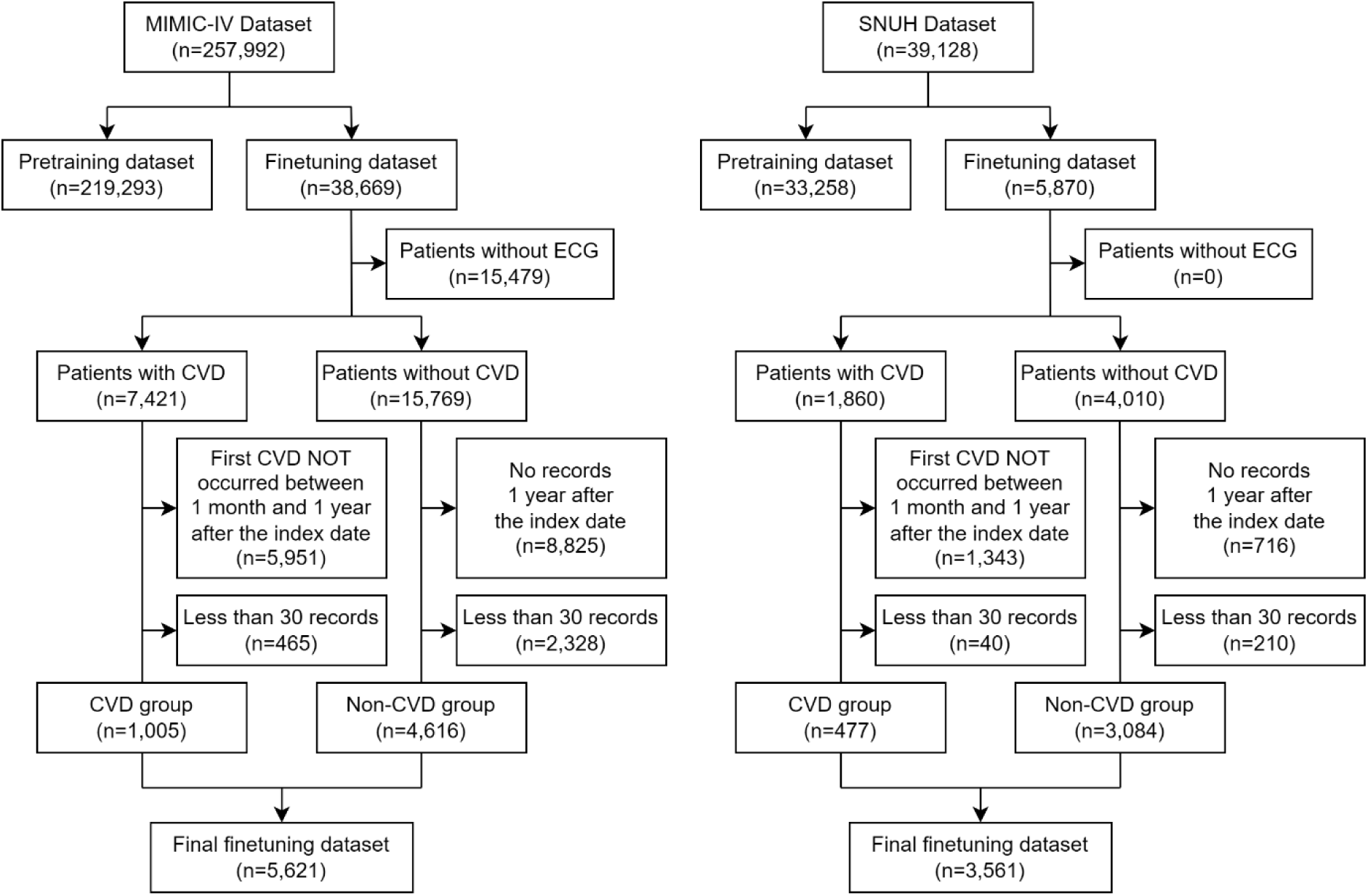
Population flowchart.

**Supplementary Table 7.** OMOP concepts for target outcomes.

| Outcome (ICD-10 codes) | OMOP concepts |
| --- | --- |
| Heart failure (I50) | 40482727, 4233424, 40480603, 439846, 4215802, 44782718, 319835, 37309625, 40481043, 4273632, 4242669, 40480602, 4229440, 4004279, 316139, 4014159, 44782719, 4195785, 44782733, 443580, 443587, 40479576, 40479192, 40481042, 444031 |
| Ischemic heart disease (I20, I21, I24) | 4270024, 4108215, 321318, 4127089, 312327, 37309626, 4121477, 44783791, 4329847, 319844, 43020460, 45766116, 4296653, 315296, 438170, 434376, 4201629, 45766075, 46270163 |
| Coronary artery disease (I25) | 36712983, 40482638, 43020480, 4119613, 4185932, 43021858, 36714444, 4127089, 321318, 4124683, 764123, 317576, 44783791, 44784217, 315286, 40481132, 36712779, 761735, 315296, 4187067, 4270354, 37118984, 4110961, 443563, 316427, 37312532, 40482655, 43021857, 40481919, 4201629, 35615052, 36712982, 314666, 438168, 37115756 |
| Stroke (I60, I61, I63) | 4148906, 4110192, 43530674, 4213731, 4274969, 4338227, 321887, 443454, 4108356, 43530727, 46273649, 313226, 4006294, 4144154, 4111708, 4108952, 4110190, 443239, 4319328, 4043731, 4110189, 4111714, 4176892, 4110186, 45767658, 4311124, 4288310, 45772786, 46270031, 42535424, 432923, 42535425, 4273526, 4110185, 4049659, 376713, 42539269 |

**Supplementary Table 8.** Example descriptions of ECG diagnoses.

| ECG diagnosis | Description |
| --- | --- |
| Normal ECG | A normal ECG indicates the heart's electrical activity is functioning within established parameters, reflecting healthy cardiac rhythm, rate, and conduction. Clinically, this suggests no significant structural or electrical abnormalities, such as arrhythmias, ischemia, or hypertrophy. Patients typically present without symptoms, but normal ECGs can also be part of routine examinations to assess cardiovascular health. Monitoring continues for risk factors like hypertension or hyperlipidemia to maintain heart health. |
| Sinus bradycardia | Sinus bradycardia is characterized by a heart rate of less than 60 beats per minute, originating from the sinus node, and is often associated with increased vagal tone, myocardial ischemia, or various medications. Clinically, patients may experience symptoms such as fatigue, dizziness, or syncope, particularly if the bradycardia is severe. Treatment involves addressing underlying causes, and if symptomatic, options may include atropine for acute relief, or pacing in cases of significant bradycardia. Regular monitoring is essential, especially in patients with underlying heart conditions or those on bradycardic medications. Long-term management may involve lifestyle modifications and evaluation for potential pacemaker implantation if symptomatic episodes persist. |
| Atrial flutter | Atrial flutter is characterized by a rapid reentrant circuit in the atria, leading to a typical "sawtooth" pattern observed on the ECG, particularly in the inferior leads (II, III, aVF), known as "F-waves." Clinically, it may present with palpitations, shortness of breath, or signs of heart failure. Treatment options include rate control using beta-blockers or calcium channel blockers, rhythm control via antiarrhythmic medications like sotalol or amiodarone, and catheter ablation for persistent cases. Anticoagulation is crucial to prevent thromboembolic events, as the stagnant blood flow in the atria increases the risk of stroke. Addressing underlying causes, such as hypertension or structural heart disease, is essential in management. |
| LVH with secondary repolarization abnormality | Left ventricular hypertrophy (LVH) often indicates underlying conditions such as hypertension or aortic stenosis, leading to the thickening of the heart's ventricular walls. The secondary repolarization abnormality suggests disturbed electrical reactivation of the ventricles, commonly manifesting as ST-segment and T-wave changes on the ECG. Clinically, patients may present with symptoms of heart failure or arrhythmias, as the heart struggles to accommodate increased workload. Treatment typically focuses on managing the underlying cause; for instance, antihypertensive medications are crucial for patients with hypertension to reduce cardiac strain. Regular monitoring and lifestyle modifications, including diet and exercise, are essential components of a comprehensive management plan. |
| Incomplete right bundle branch block | Incomplete right bundle branch block (IRBBB) reflects delayed conduction through the right bundle branch of the cardiac conduction system. Clinically, it is often associated with right ventricular hypertrophy, pulmonary hypertension, or structural heart diseases. Patients usually exhibit a normal QRS duration with characteristic ECG changes such as an rsR' pattern in lead V1. Treatment focuses on managing any underlying conditions, such as heart failure or lung disease, rather than the block itself, as IRBBB is often considered a benign finding in isolation. Regular monitoring and evaluation for progression are essential, especially if new symptoms develop. |
| Extensive ST-T changes may be due to hypertrophy and/or ischemia | Extensive ST-T changes on an ECG often indicate underlying cardiac issues, such as left ventricular hypertrophy or myocardial ischemia. In cases of hypertrophy, the heart works harder due to conditions like hypertension or valvular disorders, leading to altered repolarization patterns. Ischemia results from reduced blood flow to the myocardium, commonly caused by coronary artery disease, and manifests as ST segment depression or T wave inversions. Treatment may involve managing underlying conditions through lifestyle changes, medications such as beta-blockers or ACE inhibitors, and interventions like angioplasty or bypass surgery in ischemic cases. Prompt identification and management of these ECG changes are crucial to prevent further cardiac complications. |
| Inferior T wave changes are nonspecific | Inferior T wave changes on an ECG may indicate potential ischemia or other cardiac abnormalities, but their nonspecific nature often requires correlation with clinical symptoms and additional studies. Such changes can be seen in conditions like myocarditis, pericarditis, or electrolyte imbalances. Treatment may involve addressing underlying causes, such as revascularization procedures for ischemia or medication management for electrolyte disturbances. Continuous monitoring and follow-up ECGs are essential to determine if the changes are stable or progressive. Further investigation through cardiac imaging or stress testing may also be warranted based on the clinical scenario. |

**Supplementary Table 9.** Configuration of the model. N and L indicate the batch size and the number of ECG leads.

|  | **Network size**<br>**(kernel size, input size, output size)** | **Matrix size** |
| --- | --- | --- |
| **ECG Processing Layer** |  |  |
| Input |  | (N, lead, 5000) |
| Reduction Block1 (stride=2, padding=5, MaxPool=2) | (11, L, $8 \times L$) | (N, $8 \times L$, 1250) |
| Reduction Block2 (stride=2, padding=5, MaxPool=2) | (7, $8 \times L$) | (N, $8 \times L$, 313) |
| Reduction Block3 (stride=2, padding=5, MaxPool=2) | (5, $8 \times L$) | (N, $8 \times L$, 80) |
| Reduction Block4 (stride=1, padding=2) | (5, $8 \times L$) × 2 | (N, $8 \times L$, 80) |
| Reduction Block5 (stride=1, padding=2) | (5, $8 \times L$) × 2 | (N, $8 \times L$, 80) |
| Reduction Block6 (stride=1, padding=2, MaxPool=2) | (5, $8 \times L$, $16 \times L$) | (N, $16 \times L$, 40) |
| Reduction Block7 (stride=1, padding=2) | (5, $16 \times L$, $16 \times L$) | (N, $16 \times L$, 40) |
| Reduction Block8 (stride=1, padding=2) | (5, $16 \times L$, $16 \times L$) | (N, $16 \times L$, 40) |
| Reduction Block9 (stride=1, padding=2, MaxPool=2) | (5, $16 \times L$, $32 \times L$) | (N, $32 \times L$, 20) |
| Reduction Block10 (stride=1, padding=2) | (5, $32 \times L$, $32 \times L$) | (N, $32 \times L$, 20) |
| Reduction Block11 (stride=1, padding=2) | (5, $32 \times L$, $32 \times L$) | (N, $32 \times L$, 20) |
| **Patient Information Processing Layer** |  |  |
| Input |  | (N, 2) |
| Dense1 | (None, 2, 32) | (N, 32) |
| Dense2 | (None, 32, 64) | (N, 64) |
| Dense3 | (None, 64, 64) | (N, 64) |
| **Diabetes Detection Layer** |  |  |
| Input | | (N, $32 \times L \times 20 + 64$) |
| Dense1 | (None, $32 \times L \times 20 + 64$, 256) | (N, 256) |
| Dense2 | (None, 256, 256) | (N, 256) |
| Dense3 | (None, 256, 2) | (N, 2) |

## Notes

### Competing Interest Statement

The authors have declared no competing interest.

### Funding Statement

This research received no external funding.

### Author Declarations

The Institutional Review Board (IRB) of Seoul National University Hospital (IRB approval No. 2409-073-1570) approved the study with a waiver of informed consent, considering that our study used retrospective and observational EHR data. The approval aligns with the principles outlined in the Declaration of Helsinki, the Korean Bioethics and Safety Act (Law No. 16372), and the Human Research Protection Program-Standard Operating Procedure of Seoul National University Hospital.

